# Phenotyping of virus-specific CD8+ effector T-cells over the course of primary EBV infection and the development of PTLD in renal transplant recipients

**DOI:** 10.1101/2025.05.01.25326787

**Authors:** Kenney G. Amirkhan, Carlos van der Putten, Sophie C. Frölke, Marie J. Kersten, Ester B.M. Remmerswaal, Michiel C. van Aalderen, F. J. Bemelman

**Affiliations:** Amsterdam UMC, University of Amsterdam, Renal Transplant Unit, Department of Internal Medicine, Amsterdam Infection & Immunity, Meibergdreef 9, Amsterdam, The Netherlands; Department of Hematology, Amsterdam University Medical Centers, University of Amsterdam, Cancer Center Amsterdam, De Boelelaan 1117, Amsterdam, The Netherlands; Amsterdam UMC, University of Amsterdam, Department of Experimental Immunology, Amsterdam Infection & Immunity, Meibergdreef 9, Amsterdam, The Netherlands

**Author notes:** These authors contributed equally to this work (Kenney G. Amirkhan, Michiel C. van Aalderen, F. J. Bemelman).

## Abstract

Primary Epstein-Barr virus (EBV) infection can lead to post-transplantation lymphoproliferative disorder (PTLD) following renal transplantation (RTx). Despite EBV-driven PTLD (ePTLD) being rare, it is potentially lethal. CD8+ T-cells play a crucial role in controlling viral infections and malignant transformation of cells. We therefore hypothesized that aberrant EBV-specific CD8+ T cell maturation may contribute to ePTLD development. We performed multichannel flow cytometry on MHC class I tetramer-isolated EBV-specific CD8+ T-cells targeting lytic EBV BZLF1/BMLF1 and latent LMP2 proteins. This pilot study characterized differentiation markers, cytotoxicity, and inhibitory receptor expression in circulating EBV-specific CD8 T cells from renal transplant recipients (RTRs) with primary EBV infection and those who developed ePTLD, and compared them to RTRs with latent (inactive) EBV infection and healthy individuals (HIs). We demonstrated that RTRs with primary EBV infection display adequate cellular proliferation, development of effector memory differentiation, and expression of granzyme B and K (GrmB and K) despite immunosuppression. RTRs who developed ePTLD showed a delayed seroconversion, an early viral load peak, and altered effector-memory differentiation.

## Introduction

Epstein-Barr virus (EBV), a double-stranded DNA γ-herpesvirus, is a widespread cause of latent infection in approximately 90% of the adult immunocompetent population [1–2]. EBV is oncogenic, and under immunosuppressive circumstances, like after renal transplantation (RTx), primary infection or viral reactivation can lead to neoplastic transformation, like post-transplantation lymphoproliferative disorder (PTLD) [3]. The overall and cumulative incidence of developing PTLD after RTx is around 1-3%, with PTLD being EBV-positive in 60-80% of cases [4–5] The majority of renal transplant recipients (RTRs) ultimately (re)gain control over EBV infection, making EBV-driven PTLD (ePTLD) a rare but nevertheless potentially fatal condition, with an estimated 50-60% 5-year survival rate [6–7].

CD8+ T-cells, which have the ability to detect intracellular viral proteins, serve as an important line of defense against viruses and oncogenic transformation. Previously, our research into virus-specific CD8+ T-cells targeting polyomavirus BK (PyVBK) epitopes in RTRs revealed an impaired development of CD8+ effector-memory T cells in patients who developed PyVBK-associated interstitial nephritis when compared to RTRs who were able to control the virus [8–9]. In line with these findings, we hypothesized that variations in EBV-specific CD8+ T cell maturation are also implicated in the development of ePTLD.

To test this hypothesis, we performed a pilot study using multichannel flow cytometry on MHC class I tetramer-isolated EBV-specific CD8+ T cells. We aimed to (I) characterize the course of expression markers of differentiation, cytotoxicity, and inhibitory receptors of circulating EBV-specific CD8 T-cells in primary EBV-infected RTR patients (n = 11) and to (II) compare these characteristics to those of EBV-specific CD8+ T-cells in RTRs with latent (but inactive) EBV infection (n=9), healthy individuals (n=5), and RTRs who developed ePTLD (n=2). We phenotyped CD8+ T-cells from blood, targeting lytic EBV BZLF1/BMLF1 and latent LMP2 proteins, in which the latter is dominant in oncogenic transformation of infected cells [10–11].

## Methods

### Study group

The study population consisted of eleven RTRs, who were seronegative and PCR-negative for EBV prior the Tx and whose PBMC were extracted shortly after Tx at different time points. We extracted PBMC via standard density gradient centrifugation and cryopreserved them till the day of thawing and analysis. The demographic characteristics of our participants are shown in S1 Table. qPCR measurements were utilized to estimate viral load, whereas serological assays were employed to detect presence or absence of antibodies. Two of the 11 RTRs developed ePTLD. For comparison purposes, we included healthy individuals with latent EBV infection (n=5) and RTRs with latent EBV infection (n=9). The participants involved were enrolled in the Amsterdam UMC’s NTX BioBank. They were over the age of 18 and supplied written consent to participate. Therefore, the protocol adheres to the Declaration of Helsinki’s ethical guidelines and has been approved by the Medical Ethical Committee of the Academic Medical Center.

### Isolation of mononuclear cells from PB, flow cytometry and tetrameric complexes

PBMC were washed in PBS containing 0.01% NaN3, 2mM EDTA, and 0.5% BSA. Per two million cells, PBMC were incubated with an appropriate concentration of tetrameric complexes (S2 Table) for 30 minutes at 4°C in a dark environment while shaking, whereafter, the fluorescence-labeled monoclonal antibodies (mAbs) at concentrations according to the manufacturer’s instructions were added, followed by a further incubation of 30 min at 4°C in a dark environment while shaking. An overview of the intra- and extracellular mAbs used in this pilot study is depicted in S3 Table. A live/dead staining, Live-dead fixable RED (Invitrogen), was used to exclude dead cells from analysis. For intracellular staining a FOX-3 staining kit (Invitrogen) was used. Cells were measured on an LSR Fortessa (BD Bioscience) and analyzed with a cut-off point of a maximum of 5 cells, alongside FlowJo Version 10.10.0 software. Our gating strategy is shown in S1-S3 Figs.

### Stratification timepoints with Ki-67+ marker formula

Because the time points of PBMC collection differed between the participants, we stratified the samples based on the peak of Ki-67 expression. This consisted of a formula that detected the median of Ki-67 values, alongside the range of the median + SD * factor – median – SD * factor, with a factor value of 1 / 1.5. Afterwards, the formula was assessed by previous experiments [12–13] in order to verify the Ki-67 peak. Subsequently, we categorized the sample dates in categories, with the use of eyeballing in order for each participant to have samples in the corresponding time categories (S4 Figure).

### Statistical analysis

No statistical methods were used to predetermine sample size. Because of the relatively small group size, a non-parametric distribution was assumed, where results were displayed with the median and interquartile range (IQR) and assessed with statistical analysis using the Mann-Whitney U test, asterisks in figures indicate statistical significance (*P < 0.05, **P < 0.01, ***P < 0.001, and ****P < 0.0001). GraphPad Prism 10.2.0 was used for statistical analysis and plotting of the graphs.

## Results

### Delayed immunity and early viral load peak found in EBV patients that developed ePTLD

To synchronize the infectious stage across different RTRs with primary infection (RTRs-primary), we developed an algorithm that identifies peak Ki-67 expression, a marker of actively cycling cells, in the CD8+ T cell population. This allows us to determine the time point of maximum effector cell generation across measurements. Fig 1 depicts an overview of the course of infection in individual patients, including serum viral load, moment of seroconversion, and percentage of Ki-67 expression by total CD8+ T cells.

**Fig 1.**
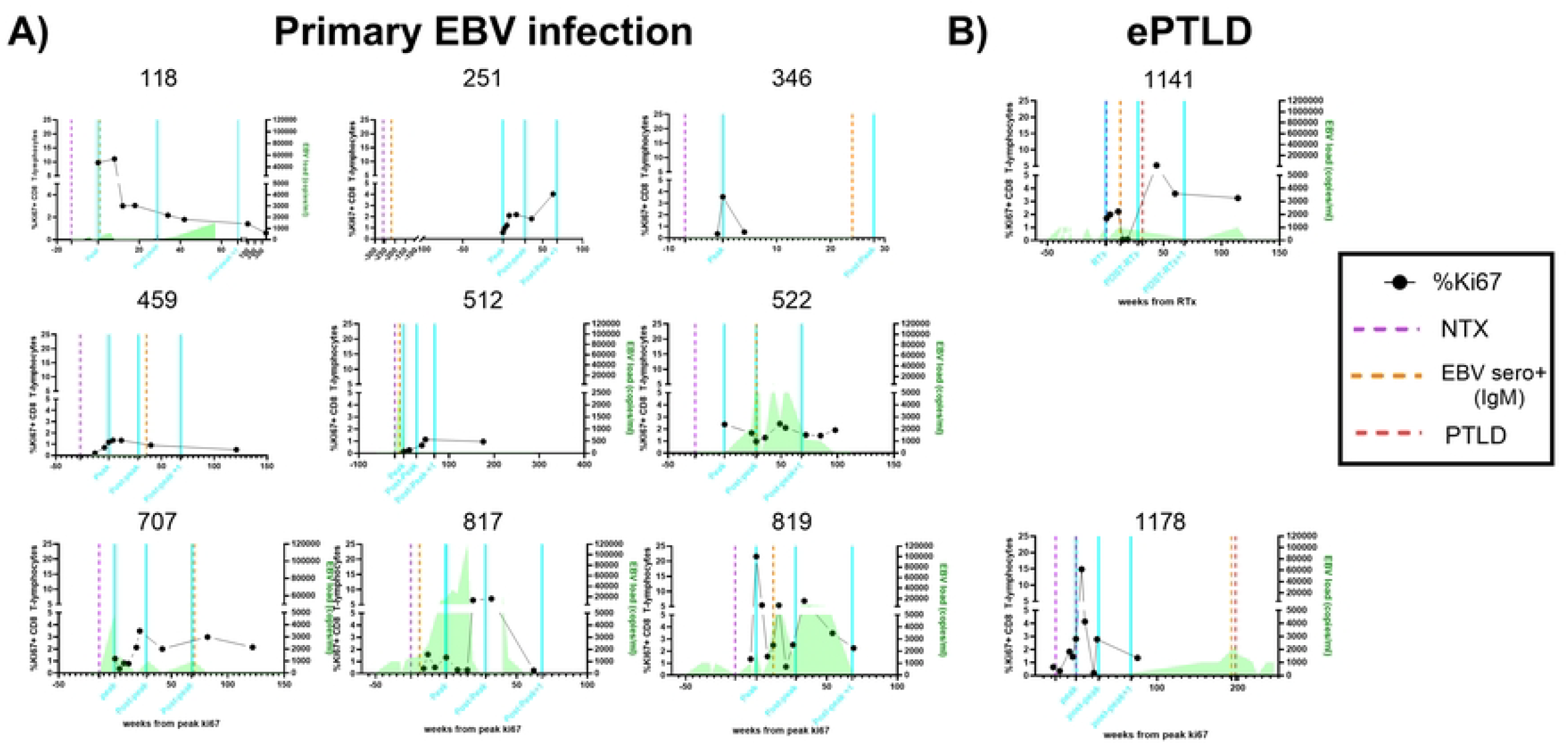
Ki67 peak of primary EBV-infected RTRs. **A)** RTRs-primary Ki67 (%) course of infection **B)** Primary EBV infected RTRs that developed ePTLD.

Among the RTRs-primary, the median time of the Ki-67 peak was detected at 20.0 weeks IQR[26.5,13.5] after RTx. The median time to seroconversion was 27 weeks after RTx IQR[12.5,85.5]. The peak EBV load (copies/ml) was detected at a median of 48.5 weeks after RTx IQR[31.5,58.5], with a median amplitude of 9345 copies/ml IQR[3668,41850]).

In the ePTLD patients, we were unable to identify a peak Ki-67 expression in the sample of patient 1141. However, in patient 1178, this occurred later when compared to the primary infection group, at 25 weeks. Also, seroconversion occurred much later with a median of 137 weeks IQR[57.0,217] after RTx. The peak viral load was detected earlier, around a median of 18.64 IQR[9.1,28.1] with a much lower median amplitude of 1630 copies/ml IQR[1000,2260].

### Dynamic shifts in EBV-specific CD8+ T cell subsets reveal unique effector memory profiles in RTRs-primary and RTRs with ePTLD

Next, we characterized the development of the effector-memory subset makeup of the EBV-specific CD8+ T cell population. Traditionally, distinct functional CD8+ T cell subsets have been defined by their surface expression of not directly related markers like CD45RA, CCR7, CD28, and CD27 [14–16], which is generally seen as a gradual process of loss and/or acquisition of surface expression of these proteins alongside epigenetic and transcriptional regulation [17–18]. Previously, using these markers, we identified the seven most prominent subsets in peripheral blood, i.e., ≥ 5% of the total CD8+ population [19]. Using multichannel flow cytometry, we phenotyped EBV-specific CD8+ T cells, targeting BZLF1- and BMLF1 epitopes (both lytic proteins) [10], as well as LMP2 epitopes, a protein directly involved in oncogenic transformation^11^, and examined their CD45RA/CCR7/CD27/CD28 expression patterns after RTx.

First, we looked at the total CD8+ T cells circulating in RTRs-primary and compared these to healthy individuals (HI). All subsets were detected to variable degrees, but HI populations contained more naïve/stem-cell-like (T_n_) cells (Fig 2A). When looking at RTRs-primary, over the course of infection, the T_n_ subset proportion decreases, and the CD45RA+ effector-memory (T_EM_RA) subset proportion increases (Fig 2B). When looking at the BZLF1/BMLF1-specific cells in HI and latently infected RTRs (RTRs-latent), the effector memory type 1 (T_EM_1) subset predominates, and only few T_EM_RA cells are seen. During primary infection, we see something similar. These cells predominantly consist of T_EM_1 cells, and their subset make-up changes only little over time with only few T_EM_RA cells. T_EM_RA cells are also barely seen among LMP2-specific populations, regardless of the study group. Here too, the T_EM_1 subset is predominating. However, during primary infection, an increase in the effector memory type 3 (T_EM_3) cells is also seen over time, a population that is rare among CD8+ T cells in HI and RTRs-latent.

**Fig 2.**
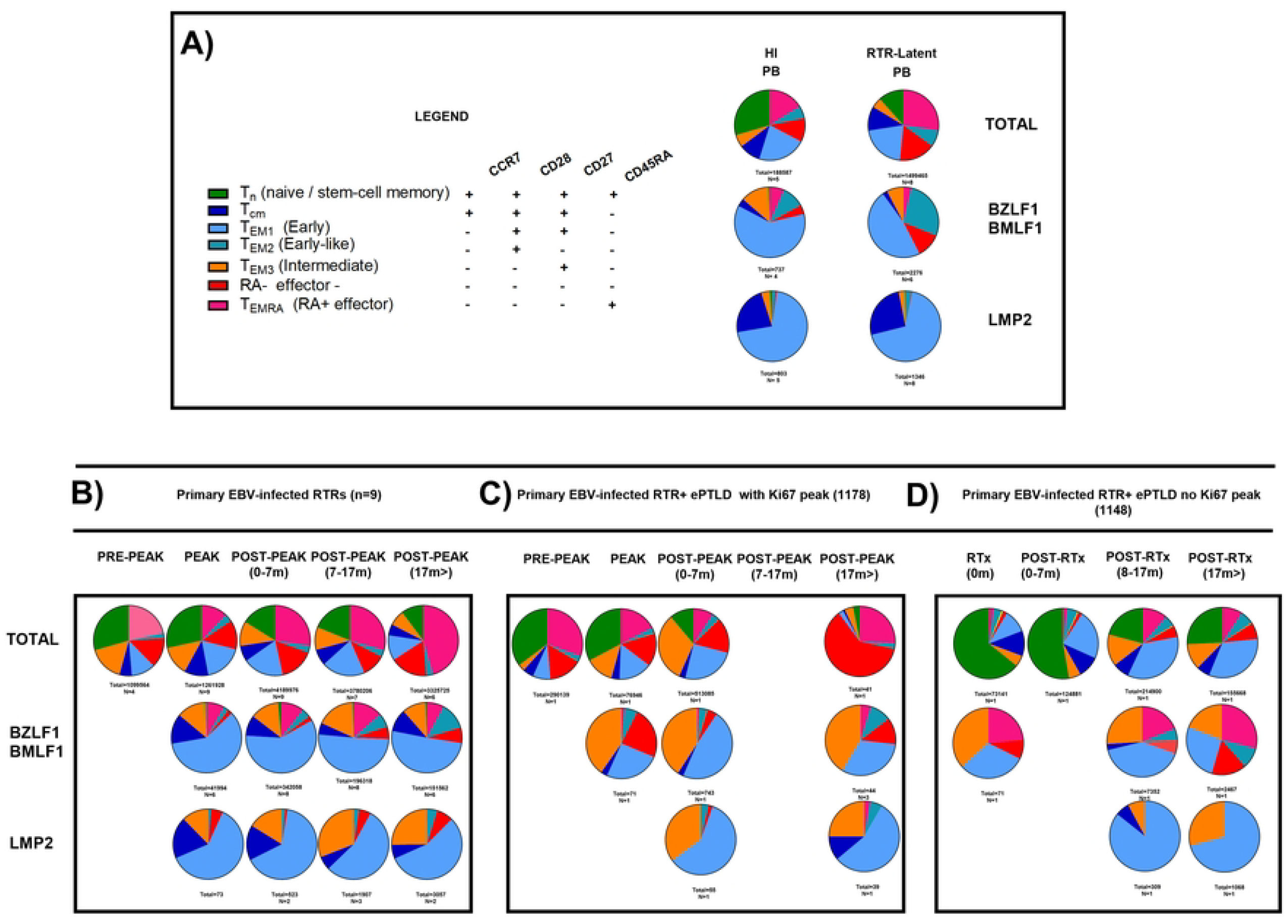
Differentiation of EBV-specific CD8 T cells in primary EBV-infected RTRs. Differentiation markers (CCR7, CD27, CD28 CD45RA-expression) for total CD8+, BZLF1/BMLF1 (lytic) and LMP2 (latent) cells. A) Immunocompetent healthy individuals (HI), RTRs-latent from peripheral blood (PB) B) RTRs-primary. C) RTR with ePTLD patient 1178 and D) RTR with ePTLD patient 1148

Next, we investigated RTRs who developed ePTLD. The total CD8+ T cell population of ePTLD patients also changed in subset make-up as time progressed. However, these changes did not seem consistent between the two individuals. When looking at the BZLF1/BMLF1-specific cells, T_EM_1 cells predominated. However, we also observed an increased proportion of T_EM_3 cells in both patients for all time points that was more pronounced than what was seen in RTRs-primary who did not develop ePTLD (Fig 2C and D). Furthermore, the central-memory (T_cm_) phenotype was less often present in comparison with RTRs-primary. When looking at the LMP2-specific populations, no substantial differences were seen between ePTLD patients and RTRs-primary.

### Divergent cytotoxic profiles and functional heterogeneity of CD8+ T cells in RTRs and RTRs with ePTLD

Next, we investigated the levels of perforin, granzyme B (GrmB), and granzyme K (GrmK) proteins in order to obtain an indirect measure of cytotoxic/antiviral functional capabilities of the T cells over the course of infection.

In the total CD8+ T cell population from RTRs-primary, we observe a significant increase in GrmB and GrmK expression as time progresses. However, levels were not statistically different from those found in HI. Interestingly, they were lower in RTRs-primary than in RTRs-latent (Fig 3A). In contrast, perforin expression levels changes were not significant, regardless of study group. When looking at the BZLF1/BMLF1 populations, the increase in GrmB was not seen. Instead, GrmB was already expressed at substantial levels from the beginning, and levels appeared to stay more or less stable over time. No differences were noted between study groups. GrmK levels showed an increasing trend without differences between study groups (Fig 3B). Perforin levels were particularly high during the pre-peak time point but decreased as time progressed. Also here, no differences were noted between study groups, besides high expression pre-peak in RTRs-primary. LMP2-specific CD8+ T cells differed from the total CD8+ and the BZLF1/BMLF1-specific population in the sense of a particularly high expression of GrmK, which was higher than what was seen for RTRs-latent but similar to HI. Also here, perforin was detected primarily during the pre-peak period, after which it declined to low levels on later time points. (Fig 3C).

**Fig 3.**
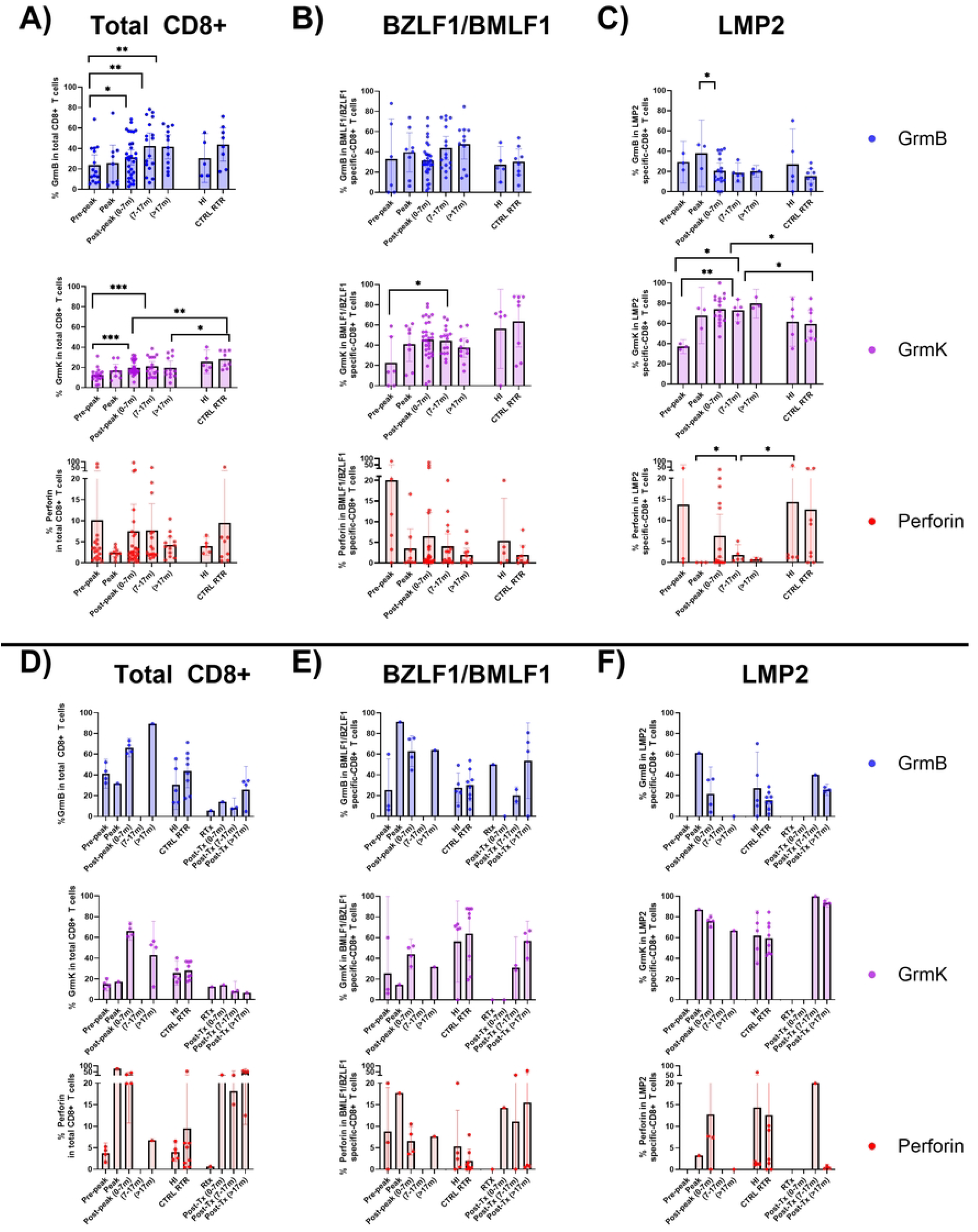
Expression of granzyme-B, and -K, and Perforin of EBV-specific CD8 T cells in primary EBV-infected RTRs (n=9) (A,B,C) and 2 RTRs with ePTLD (with Ki67 peak (Left) and without Ki67 peak (right) (D,E,F). Cytotoxicity (Granzyme B, Granzyme K, Perforin). For total CD8+, BZLF1/BMLF1 (lytic) and LMP2 (latent) cells. Healthy individuals (HI), n=5, and RTRs with latent EBV (CTRL RTR), n=9, are both from PB.

For the ePTLD patients, we observed a trend towards higher expression of GrmB among the overall CD8+ T cell population on nearly all the time points. Nevertheless, due to the low number of patients, statistical comparison was not possible. An increased expression of GrmK was also seen, but only after the Ki-67 peak. Perforin levels also increased sharply from the moment of peak Ki-67 expression (Fig 3D). BZLF1/BMLF1 cells displayed higher levels of GrmB on the peak and post-peak time points when compared to RTRs-primary. In contrast, for GrmK, the levels were lower at these time points (Fig 3E). When looking at the LMP2-specific cells, GrmB levels were increased at the Ki-67 peak when compared to RTRs-primary. In addition, particularly high levels of GrmK and perforin expression were also noted for all time points (Fig 3F).

In addition, we also analyzed different phenotypes of cytotoxic expression based on combined expression of GrmB, GrmK, and perforin (S5 Fig). The triple expression phenotype is present in LMP2-specific CD8+ T cells in controls, RTRs-primary and decreases over time (S5B Fig). This phenotype is also present in one RTR with ePTLD in every analyzed EBV-specific CD8+ T-cells, whereas in the other RTR with ePTLD this phenotype is mostly absent in all analyzed cells.

### Contrasting inhibitory dynamics in EBV-specific CD8+ T cells differentiate ePTLD from normal immune response

Next, we wanted to know to what extent co-inhibitory receptors, markers of T-cell exhaustion, are implicated in the normal anti-EBV response and whether differences could be seen for T cells in RTRs who developed ePTLD. Inhibitory receptors, such as PD1, CD160, and CD244, have indeed previously been described to play a pivotal role in later stages of chronic viral infections [20–21].

When looking at the total CD8+ T cells in RTRs-primary, we observe an increase in CD160, CD244, and PD1 expression, over the course of infection (Fig 4A). However, expression levels did not differ drastically from HI and RTRs-latent. BZLF1/BMLF1-specific cells show high expression levels of all three inhibitory receptors in comparison with total CD8+ T cells, which further increase over time for CD160 and CD244. However, these levels were generally not different from those found in HI and RTRs-latent (Fig 4B). LMP2-specific cells in RTRs-primary showed a high expression of CD160, particularly after the Ki-67 peak, which was higher than the levels found in HI and RTRs-latent. CD244 levels were high at all time points but did not differ from the levels of HI and only differed significantly at post-peak with RTRs-latent. PD1 levels were also high at all time points, being increased when compared to HI and RTRs-latent. (Fig 4C).

**Fig 4.**
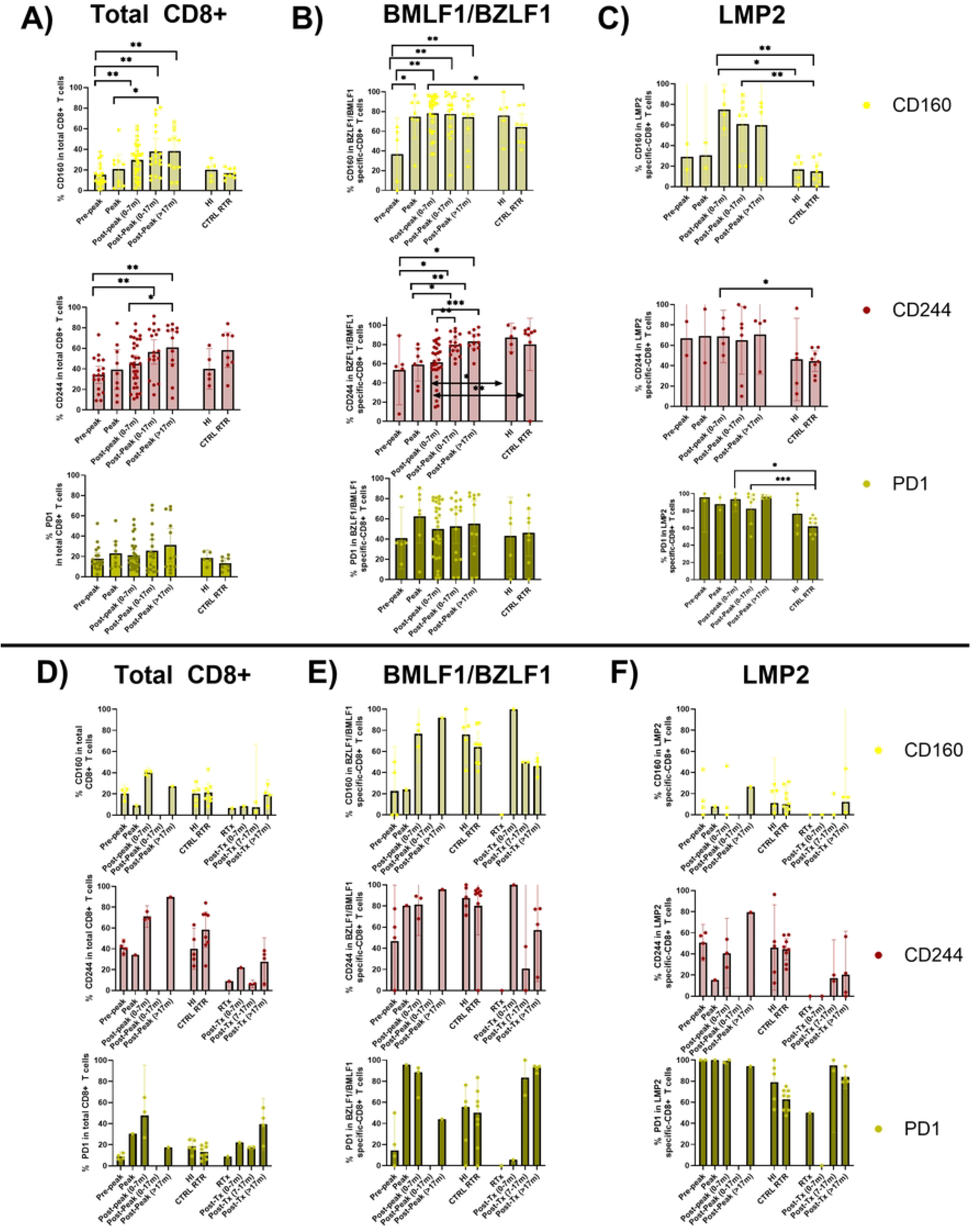
Expression of inhibitory receptors, CD160, CD244, and PD1 of EBV-specific CD8 T cells in primary EBV-infected RTRs (n=9) (A, B, C) and 2 RTRs with ePTLD (with Ki67 peak (left) and without Ki67 peak (right)) (D,E,F). For total CD8+, BZLF1/BMLF1 (lytic) and LMP2 (latent) cells. Healthy individuals (HI), n – 5, and RTRs with latent EBV, n = 9 (CTRL RTR) both from PB.

When looking at the ePTLD patients, CD160, CD244 and PD1 levels did not appear to differ much from what was seen in the other study groups (Fig 4D-F).

Besides their inhibitory effects, combined expression of CD160 and CD244 was reported to also confer cytolytic potential [22]. Therefore, we also analyzed combinatorial expression of these expression markers (CD160, CD244, and PD1) (S6 Fig). We observed a high level of CD244+CD160+PD1+ triple expression by the LMP2-specific cells in RTRs-primary subjects compared to other study groups (S6B Fig).

## Discussion

In the current manuscript, we describe phenotypical changes in relation to viral load over the course of primary EBV infection in RTRs. In addition, we provide limited but rare data on primary infection prior to ePTLD development in two RTRs. We found that RTRs who developed ePTLD showed a delayed seroconversion, an early viral load peak, and altered effector-memory differentiation.

In addition, for one of the RTRs with ePTLD, no Ki-67 peak was found. Despite the fact that we were able to identify a Ki-67 peak in all other subjects, we acknowledge that the limited number of PTLD patients and non-standardized sampling moments may be a factor. Nevertheless, the underlying mechanism for a lack of a Ki-67 peak may involve T-cell exhaustion, a process where T cells have altered activation and differentiation, in the presence of persistent antigen exposure [23]. Proteins in the EBV genome exhibit immune evasion tactics that can also affect the strength of the T cell response, particularly against a background of immunosuppressive medication [24]. Furthermore, we observed that the Ki-67 peak for the other subject who developed ePTLD occurred a month later than for the RTRs with primary infection who did not develop ePTLD, further supporting the hypothesis of a diminished T cell proliferation as a factor involved in the pathogenesis of ePTLD.

Within the overall CD8+ T-cell population, we observed a decrease in naïve T cells and an increase in T_EM_RA cells, as expected. This fits a model where naïve T cells are recruited to terminally differentiate into T_EM_RA cells, cells that normally possess strong cytolytic potential [25]. Interestingly, this increase of T_EM_RA cells was not seen in the BZLF1/BMLF1 and LMP2-specific populations; therefore, in future studies, more virus-specific populations targeting other EBV proteins need to be assessed.

In addition, a substantial proportion of the normally also cytotoxic T_EM_3 subset was identified in the developing virus-specific T cell populations, particularly concerning the BZLF1/BMLF1-specific cells in the ePTLD patients. Whereas T_EM_RA cells predominantly express GrmB, the T_EM_3 cells are known for their high concomitant expression of GrmB and GrmK (as opposed to the T_cm_, T_EM_1 and T_EM_2 subsets that are generally not carrying these molecules) [19]. Granzyme B is known to initiate apoptosis after injection into an infected cell via a pore created by perforin. On the other hand, studies have demonstrated that GrmK directly targets viruses within cells. A study identified a distinct subset of GrmK-expressing CD8 T cells that modulate neutrophilic inflammation in nasal polyps that were associated with EBV infection and do not express GrmB nor perforin [26]. This suggests a perforin-independent manner of GrmK, GrmK can extracellularly trigger pro-inflammatory cytokines and stimulate inflammation and recruitment of activated immune cells [27–28]. However, it is still unknown how GrmK can specifically enter the cell, independent of perforin. A possible mechanism may involve GrmK-mediated inhibition of important pathways of viral protein replication and assembly, as it was found to block nucleoprotein (NP) and viral RNA complex formation into the host nucleus in vitro, allowing for viral clearance during influenza infection [29].

Interestingly, in the BZLF1/BMLF1 population, GrmB expression predominated over GrmK, while in LMP2-specific cells, GrmK predominated over GrmB, as expected [13]. However, in these cells, we observe low amounts of perforin in the presence of GrmK. This suggests a different functional requirement of T cells when faced with different viral proteins (expressed in different phases of viral proliferation). LMP2-specific cells in RTRs without PTLD had decreasing perforin expression after Ki67 peak, with similar high GrmK expression and lower GrmB expression, when compared to RTRs with ePTLD. RTRs with ePTLD show high perforin expression and high GrmB and K expression. This indicates incorrect skewing causing these patient’s immune response to diverge.

We finally observed high and increasing expression levels of co-inhibitory receptors CD160, CD244, and PD1 in RTRs with primary or latent infection. Despite our findings on an absent Ki-67 peak in one of RTRs who developed ePTLD, no obvious differences were noted for the PTLD patients with regard to the expression of co-inhibitory receptors, but the small size of the group remains an important limitation of the study. Previous studies have shown that inhibitory receptor expression, like PD1, is necessary for tolerogenic purposes to gain control over EBV, suggesting that expression of co-inhibitory receptors is indeed beneficial for primary EBV infection [30]. Also, to truly assess function inhibition and/or T cell exhaustion, further functional experiments that examine actual in vitro killing and additional cytokine production are required in future studies.

This pilot study provided insights into the phenotypical changes of virus-specific CD8+ T cells in RTRs during primary EBV infection. We phenotyped RTRs suffering from primary EBV infection, based on differentiation, cytotoxicity, and inhibitory receptor markers, which showed sufficient cellular proliferation, development of the T_EM_3 population, and expression of GrmK even under immunosuppressive circumstances. To further understand how ePTLD develops in RTRs and how aberrancies in the proliferation and differentiation of antiviral CD8+ T cells may be implicated in the pathogenesis of ePTLD, more research on a larger population of subjects developing ePTLD is needed.

## Data Availability

All relevant data are within the manuscript and its Supporting Information files.The data that support the findings of this study are available from the corresponding author, upon reasonable request.

